# Incidence of Long COVID Following Reinfection with COVID-19

**DOI:** 10.1101/2025.08.12.25333155

**Authors:** M Daniel Brannock, Emily Hadley, Alexander Preiss, Megan L Fitzgerald, Nita Jain, Emily Taylor, Andrew Wylam, Yun J Yoo, Elaine Hill, Richard A Moffitt, the N3C and RECOVER consortia

**Affiliations:** RTI International, RTP, NC; Patient-led Research Collaborative; Timeless Biosciences, Atlanta, GA; Solve M.E., Los Angeles, CA; Pandemic Patients; Emory University, Atlanta, GA; University of Rochester, Rochester, NY

## Abstract

**Background:** COVID-19 reinfections have emerged as a critical concern, particularly in relation to post-acute sequelae of SARS-CoV-2 infection, commonly known as long COVID. Long COVID is known to manifest diverse, debilitating symptoms across all demographics. Limited studies have investigated the causal relationship of COVID-19 reinfections and long COVID.

**Methods:** We leveraged demographically diverse electronic health records from the COVID-19 enclave of the National Clinical Cohort Collaborative, part of the RECOVER initiative, to create a matched cohort of reinfected and control adults. All participants had at least one documented COVID-19 infection. We used one-to-one coarsened exact matching on sex, race/ethnicity, age, healthcare utilization, existing comorbidities, site of care, and the timing and severity of first infection. Index dates were assigned to each matched pair as the date of reinfection for the reinfected case. Long COVID was defined using a machine learning computable phenotype trained on clinically diagnosed long COVID cases. Cumulative incidence one year after index was calculated using an Aalen-Johansen estimator. Risk ratios were calculated by taking the ratio of long COVID incidence among reinfected and control cases.

**Results:** We found that reinfection resulted in a significantly higher risk of long COVID compared to not being reinfected (risk ratio, 1.35, 95% CI, 1.32-1.39; risk difference, 0.029, 95% CI, 0.027-0.031). This effect was consistent across most stratifications.

**Conclusions:** We found that COVID-19 reinfection resulted in a roughly 35% increase in the incidence of long COVID in a matched cohort using observational electronic health records.

---

It is well documented that individuals can experience multiple infections with SARS-CoV-2, or COVID-19 reinfections.^1–3^ A potentially debilitating outcome of COVID-19 can be post-acute sequelae of SARS-CoV-2 infection, also known as long COVID.^4,5^ In this study, we use a robust matching methodology and a large cohort of patients in the electronic health record (EHR)- based National Clinical Cohort Collaborative (N3C) Data Enclave to assess the impact of COVID-19 reinfections on long COVID.

Reinfections may occur due to waning immunity from prior vaccination or COVID-19 infection.^2^ Prior research has established some characteristics of reinfections, including their incidence and severity. ^6–8^ COVID-19 reinfections are common and widespread ^3,9,10^. Experiencing a more severe first infection may increase the likelihood of a more severe reinfection requiring hospitalization.^6^ Research on COVID-19 vaccinations and reinfections has been contradictory, with Butt et. al suggesting that vaccination was protective against hospitalization or death following a reinfection while Bowe et. al suggests that there was no difference in likelihood of adverse events like hospitalization following reinfection by vaccination status.^7,9^

Long COVID is a post-COVID condition with a variety of symptoms that negatively impact health and well-being.^5,11^ Patient symptom cluster analysis has suggested that symptoms can be wide-ranging, spanning cognitive, pulmonary, and cardiovascular body systems.^12–14^ Long COVID affects individuals of all ages and demographics, with global incidence estimated at more than 400 million individuals in 2023.^5,15–17^ Evidence suggests that COVID-19 vaccination may reduce the risk of long COVID.^18–20^ Long COVID is chronic and has no cure, requiring ongoing maintenance.^4,11^

Limited research has explored COVID-19 reinfections and long COVID. There is agreement that de novo long COVID may occur following a reinfection.^5,21^ Bowe et. al studied 5.8 million patients in the US Department of Veterans Affairs’ national healthcare database and found that the cumulative risk of long COVID increased with each reinfection.^7^ An online survey found that COVID-19 reinfections are associated with increased odds of reporting long COVID, and that reinfections worsen long COVID symptoms.^22^ However, Bosworth et. al found that among more than 300K participants in the UK COVID-19 infection survey, the risk of de novo long COVID after a second SARS-CoV-2 infection was lower than after a first infection.^23^ Researchers have highlighted the limited research and called for increased study of the relationship between reinfections and long COVID when considering vaccination and prior infections.^21^

In this study, we address these gaps by first proposing a more comprehensive methodology using one-to-one coarsened exact matching. We also include a subanalysis that considers how vaccination status modulates the effect of reinfection. Finally, we discuss broader implications and opportunities for future work.

## Methods

Our data came from the COVID enclave of the National Clinical Cohort Collaborative (N3C), a part of the NIH Researching COVID to Enhance Recovery (RECOVER) Initiative.^24^ It includes electronic health records (EHRs) from 23 million individuals in the United States dating back to January 1, 2018 from 85 hospitals and hospital networks. Of those individuals, nearly 9 million have documented cases of COVID-19. We used N3C release version 187, dated December 13, 2024, and a subset of 64 sites that met data quality standards. All data analysis was performed in Palantir Foundry (2021, Denver, CO).

### IRB

Data transfer in N3C is performed under Johns Hopkins University Reliance Protocol # IRB00249128 or individual site agreements with NIH. N3C is managed under the authority of the NIH. For more information see https://ncats.nih.gov/n3c/resources.

### Definitions

#### Inclusion and exclusion criteria

All individuals in our study had a documented COVID-19 infection between March 1, 2020 and December 4, 2023, as evidenced by a U07.1 diagnosis, positive SARS-CoV-2 PCR or antigen test, or a nirmatrelvir-ritonavir prescription; had a documented sex; were between 18 and 99 years old at the time of their first documented COVID-19 infection; and did not have long COVID prior to their first documented COVID-19 infection.

#### Outcome definition

Our outcome was a long COVID computable phenotype as defined by a machine learning model.^25,26^ Individuals with symptoms similar to those observed in clinically diagnosed cases of long COVID were labeled with long COVID. A detailed description of how the machine learning model was used to define long COVID is included in the supplemental materials.

#### Reinfection definition

COVID-19 reinfections were defined by a positive SARS-CoV-2 PCR or antigen test occurring at least 60 days after a previously documented infection.^6^ We chose 60 days based on when patients stop shedding SARS-CoV-2 and with the support of the RECOVER clinician advisory panel.^27,28^ Only reinfections in the Omicron era (on or after November 1, 2021 and until the study end date of June 1, 2024) were considered. Individuals labeled as **non-reinfected** do not have a recorded reinfection at the time of their index date.

### Matching

We modified traditional coarsened exact matching (CEM)^29^ to define our cohort of reinfected and non-reinfected individuals. Individuals were matched according to their demographics and health histories. A detailed summary of the matching procedure is included in the statistical analysis plan.

### Statistical Analysis

Cumulative incidence was estimated for reinfected and non-reinfected cases using an Aalen-Johansen estimator. The relative risk of long COVID was calculated as the ratio of the cumulative incidence 365 days after the index among reinfected cases versus non-reinfected cases (the average treatment effected among “treated”, or reinfected, individuals). Individuals were censored for subsequent reinfections, loss to follow-up, study end, or death; see statistical analysis plan for details. The absolute risk difference was calculated as the difference in cumulative incidence. We estimated two-sided 95% confidence intervals from 500 bootstrap iterations. We calculated the overall relative and absolute risk and stratified by each matching criterion.

### Negative Control Outcomes

We identified two negative control outcomes^30^ to assess the robustness of our findings: administration of a Papanicolaou test (Pap test) and non-pathological bone fracture diagnosis. Both were chosen as outcomes that are unrelated to SARS-CoV-2 reinfection or long COVID onset with guidance from clinicians and long COVID patient advocates. The procedure in our primary analysis was repeated using each of the negative control outcomes, except that the Pap test cohort was limited to females aged 30 to 65 at the time of their first COVID-19 event.

### Vaccine Subanalysis

We performed a subanalysis using sites of care with robust vaccination reporting to assess how vaccination status modulates the effect of reinfections on long COVID. We identified 13 sites that had reported vaccination rates within two-thirds of CDC-reported vaccination rates in their catchment area.^18^ We repeated our analysis using only individuals from these 13 sites. We added vaccination status (having at least a single vaccination or not) prior to first COVID-19 event as a matching criterion. After pairs were assigned, we kept only those who were both vaccinated or both not vaccinated between first infection and their index date. Relative and absolute risks were calculated as in the primary analysis, with vaccination status added as a risk stratification.

## Results

Our final cohort consisted of 424,616 individuals: 212,313 pairs of reinfected and non-reinfected cases. The cohort is summarized in Table 1. Table 2 shows attrition for cohort selection.

**Table 1.**
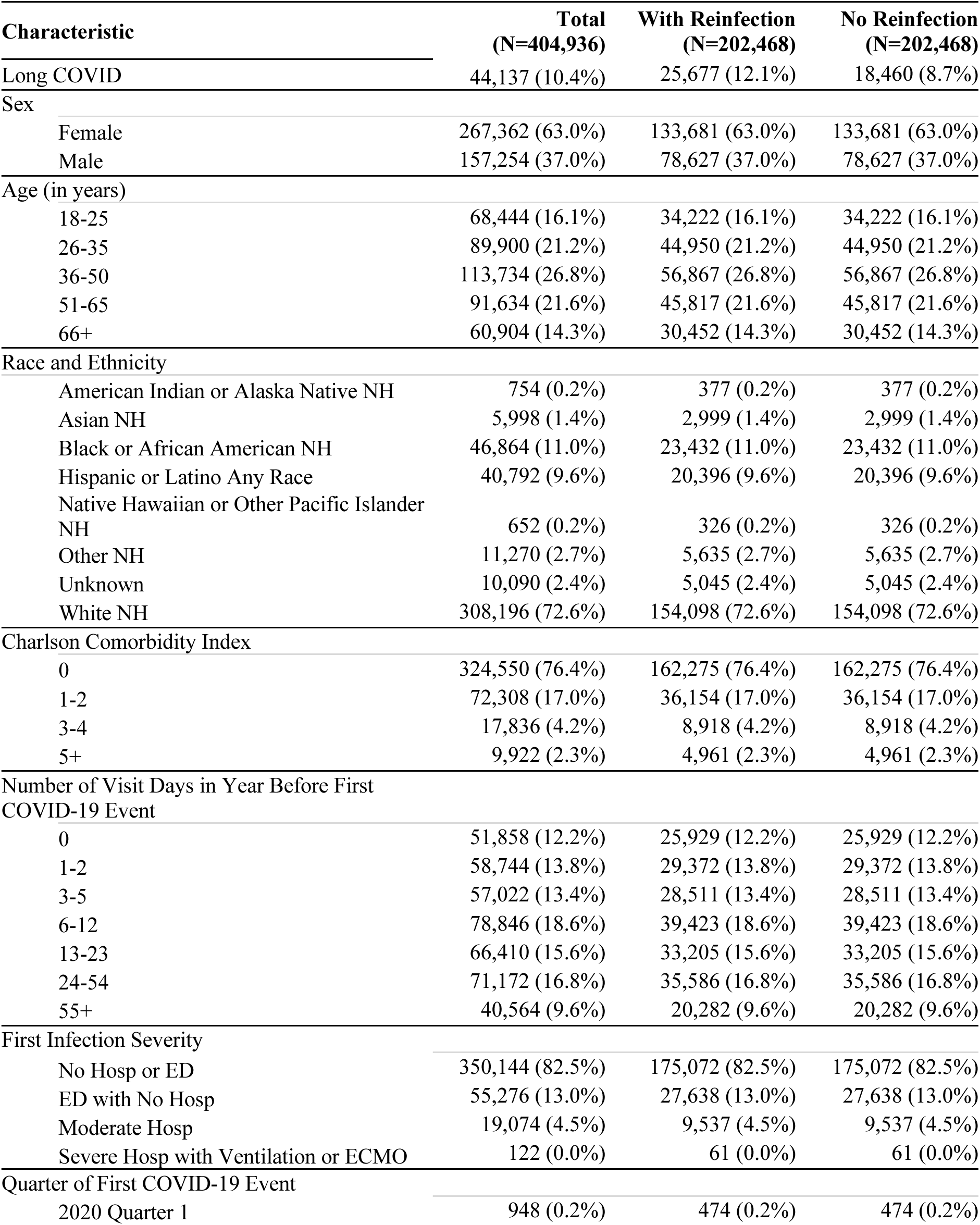
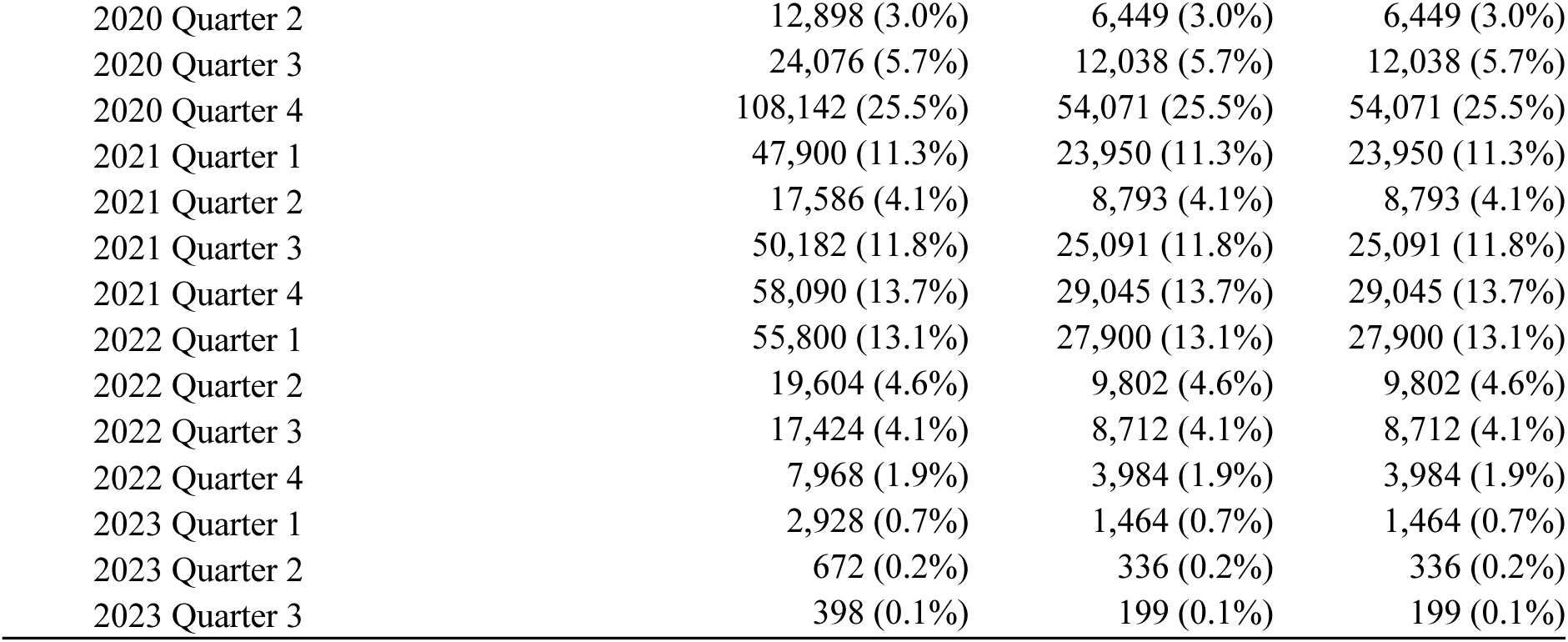
Matched cohort description. NH: Non-Hispanic, ED: Emergency Department, Hosp: Hospitalization.

| Characteristic | Total<br>(N=404,936) | With Reinfection<br>(N=202,468) | No Reinfection<br>(N=202,468) |
| --- | --- | --- | --- |
| Long COVID | 44,137 (10.4%) | 25,677 (12.1%) | 18,460 (8.7%) |
| Sex |  |  |  |
| Female | 267,362 (63.0%) | 133,681 (63.0%) | 133,681 (63.0%) |
| Male | 157,254 (37.0%) | 78,627 (37.0%) | 78,627 (37.0%) |
| Age (in years) |  |  |  |
| 18-25 | 68,444 (16.1%) | 34,222 (16.1%) | 34,222 (16.1%) |
| 26-35 | 89,900 (21.2%) | 44,950 (21.2%) | 44,950 (21.2%) |
| 36-50 | 113,734 (26.8%) | 56,867 (26.8%) | 56,867 (26.8%) |
| 51-65 | 91,634 (21.6%) | 45,817 (21.6%) | 45,817 (21.6%) |
| 66+ | 60,904 (14.3%) | 30,452 (14.3%) | 30,452 (14.3%) |
| Race and Ethnicity |  |  |  |
| American Indian or Alaska Native NH | 754 (0.2%) | 377 (0.2%) | 377 (0.2%) |
| Asian NH | 5,998 (1.4%) | 2,999 (1.4%) | 2,999 (1.4%) |
| Black or African American NH | 46,864 (11.0%) | 23,432 (11.0%) | 23,432 (11.0%) |
| Hispanic or Latino Any Race | 40,792 (9.6%) | 20,396 (9.6%) | 20,396 (9.6%) |
| Native Hawaiian or Other Pacific Islander NH | 652 (0.2%) | 326 (0.2%) | 326 (0.2%) |
| Other NH | 11,270 (2.7%) | 5,635 (2.7%) | 5,635 (2.7%) |
| Unknown | 10,090 (2.4%) | 5,045 (2.4%) | 5,045 (2.4%) |
| White NH | 308,196 (72.6%) | 154,098 (72.6%) | 154,098 (72.6%) |
| Charlson Comorbidity Index |  |  |  |
| 0 | 324,550 (76.4%) | 162,275 (76.4%) | 162,275 (76.4%) |
| 1-2 | 72,308 (17.0%) | 36,154 (17.0%) | 36,154 (17.0%) |
| 3-4 | 17,836 (4.2%) | 8,918 (4.2%) | 8,918 (4.2%) |
| 5+ | 9,922 (2.3%) | 4,961 (2.3%) | 4,961 (2.3%) |
| Number of Visit Days in Year Before First COVID-19 Event |  |  |  |
| 0 | 51,858 (12.2%) | 25,929 (12.2%) | 25,929 (12.2%) |
| 1-2 | 58,744 (13.8%) | 29,372 (13.8%) | 29,372 (13.8%) |
| 3-5 | 57,022 (13.4%) | 28,511 (13.4%) | 28,511 (13.4%) |
| 6-12 | 78,846 (18.6%) | 39,423 (18.6%) | 39,423 (18.6%) |
| 13-23 | 66,410 (15.6%) | 33,205 (15.6%) | 33,205 (15.6%) |
| 24-54 | 71,172 (16.8%) | 35,586 (16.8%) | 35,586 (16.8%) |
| 55+ | 40,564 (9.6%) | 20,282 (9.6%) | 20,282 (9.6%) |
| First Infection Severity |  |  |  |
| No Hosp or ED | 350,144 (82.5%) | 175,072 (82.5%) | 175,072 (82.5%) |
| ED with No Hosp | 55,276 (13.0%) | 27,638 (13.0%) | 27,638 (13.0%) |
| Moderate Hosp | 19,074 (4.5%) | 9,537 (4.5%) | 9,537 (4.5%) |
| Severe Hosp with Ventilation or ECMO | 122 (0.0%) | 61 (0.0%) | 61 (0.0%) |
| Quarter of First COVID-19 Event |  |  |  |
| 2020 Quarter 1 | 948 (0.2%) | 474 (0.2%) | 474 (0.2%) |
| 2020 Quarter 2 | 12,898 (3.0%) | 6,449 (3.0%) | 6,449 (3.0%) |
| 2020 Quarter 3 | 24,076 (5.7%) | 12,038 (5.7%) | 12,038 (5.7%) |
| 2020 Quarter 4 | 108,142 (25.5%) | 54,071 (25.5%) | 54,071 (25.5%) |
| 2021 Quarter 1 | 47,900 (11.3%) | 23,950 (11.3%) | 23,950 (11.3%) |
| 2021 Quarter 2 | 17,586 (4.1%) | 8,793 (4.1%) | 8,793 (4.1%) |
| 2021 Quarter 3 | 50,182 (11.8%) | 25,091 (11.8%) | 25,091 (11.8%) |
| 2021 Quarter 4 | 58,090 (13.7%) | 29,045 (13.7%) | 29,045 (13.7%) |
| 2022 Quarter 1 | 55,800 (13.1%) | 27,900 (13.1%) | 27,900 (13.1%) |
| 2022 Quarter 2 | 19,604 (4.6%) | 9,802 (4.6%) | 9,802 (4.6%) |
| 2022 Quarter 3 | 17,424 (4.1%) | 8,712 (4.1%) | 8,712 (4.1%) |
| 2022 Quarter 4 | 7,968 (1.9%) | 3,984 (1.9%) | 3,984 (1.9%) |
| 2023 Quarter 1 | 2,928 (0.7%) | 1,464 (0.7%) | 1,464 (0.7%) |
| 2023 Quarter 2 | 672 (0.2%) | 336 (0.2%) | 336 (0.2%) |
| 2023 Quarter 3 | 398 (0.1%) | 199 (0.1%) | 199 (0.1%) |

**Table 2.**
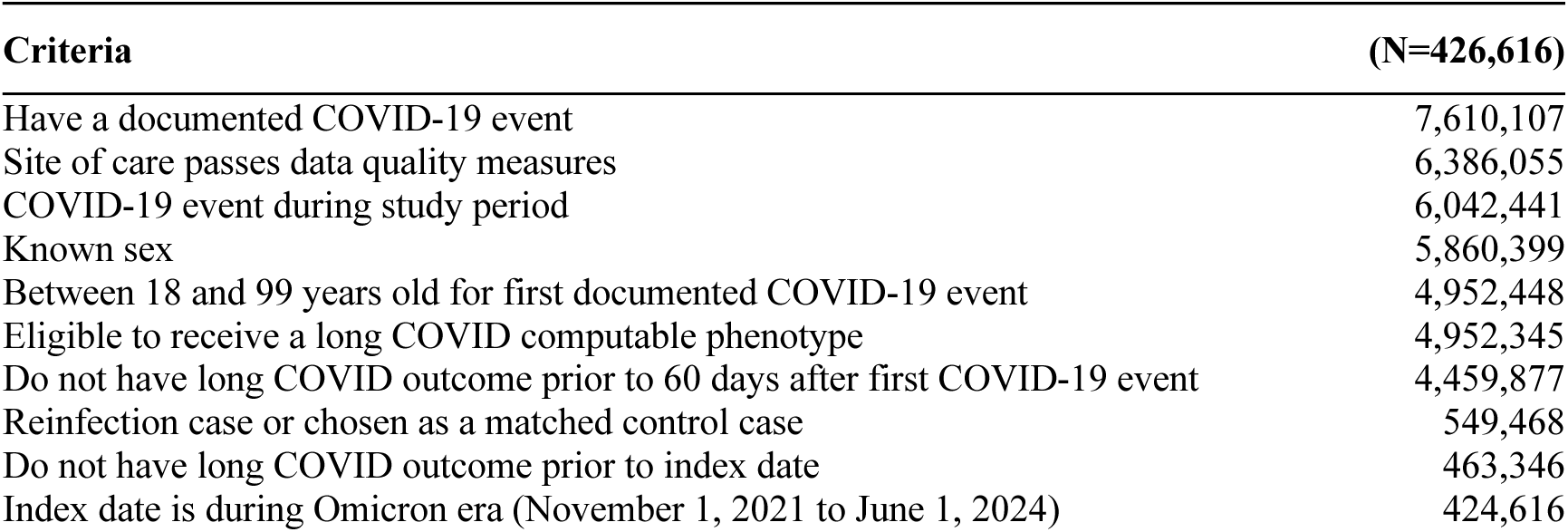
Attrition table.

There were 25,677 (12.1%) cases of long COVID among reinfected cases and 18,460 (8.7%) cases of long COVID among non-reinfected cases. After accounting for censoring, the cumulative incidence of long COVID among the reinfected and non-reinfected cases was 11.1% and 8.2%, respectively. Among patients similar to those with reinfections, we found that reinfections resulted in a significant increase in risk of developing long COVID (risk ratio, 1.35, 95% CI, 1.32-1.39; absolute risk difference, 0.029, 95% CI, 0.027-0.031). Figure 1 highlights the relative risk in our primary analysis and negative control conditions, with stratifications by age and vaccination status from the vaccination subanalysis. Figure 2 shows the cumulative incidence over time of reinfected and non-reinfected cases. Table 3 includes the cumulative incidence, risk ratio, and absolute risk difference for all strata (site of care included in eTable 1).

**Figure 1.**
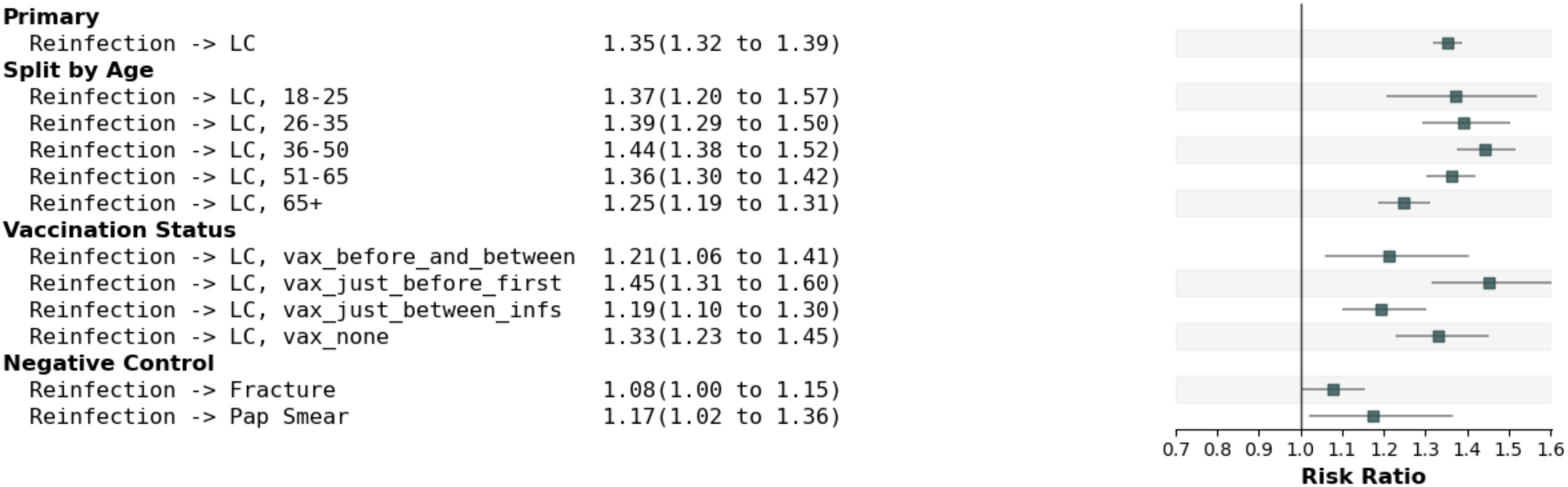
Primary analysis, vaccination analysis, select stratifications, and negative control relative risk ratios.

**Figure 2.**
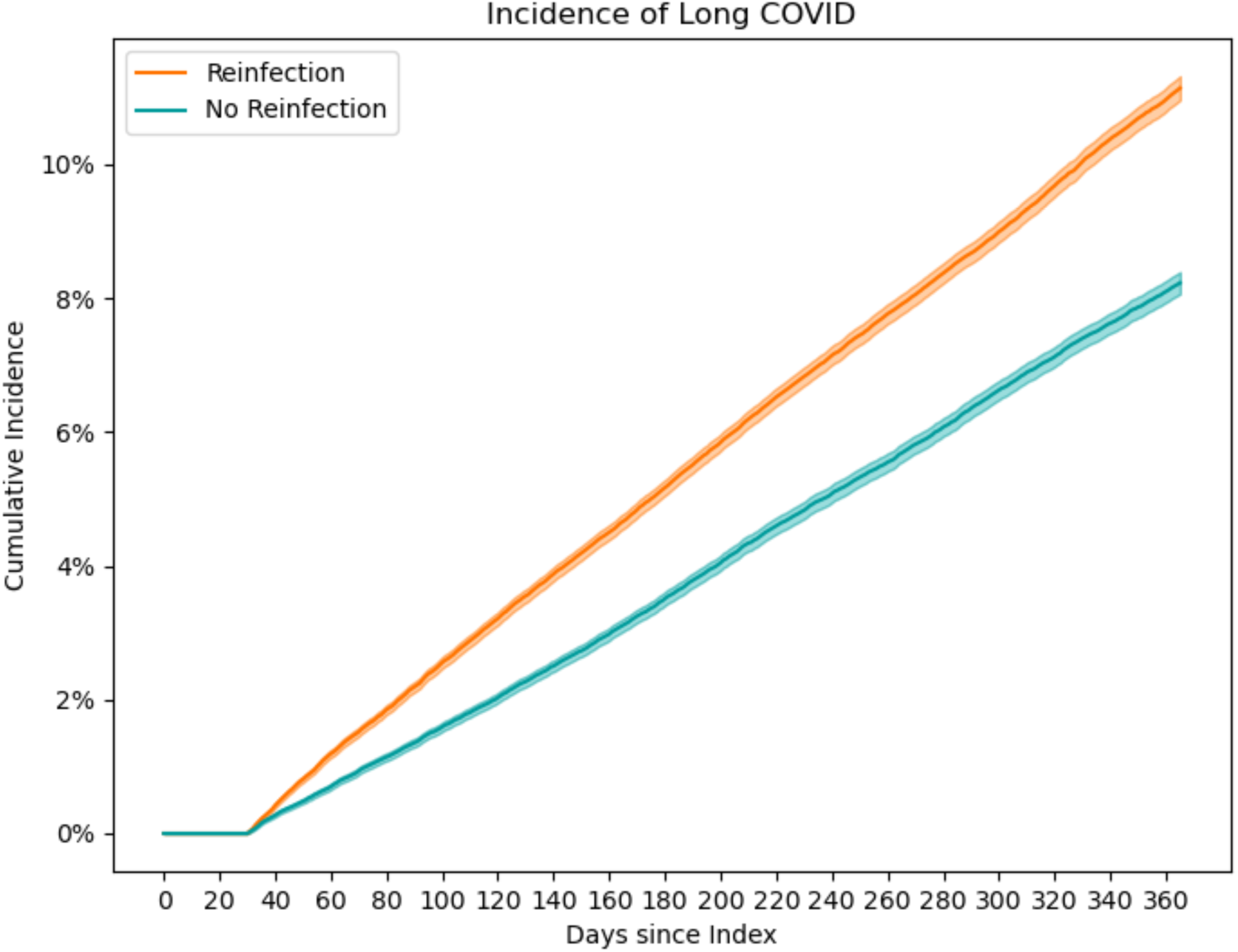
Cumulative incidence curves for reinfections and control cases. Outcomes are not allowed prior to 30 days after index, as these symptoms may be attributable to the acute reinfection.

**Table 3.** Cumulative incidence, risk ratios, and absolute risk differences for primary analysis, vaccination analysis (using the vaccination analysis cohort), negative control conditions, and stratified analysis.

| Analysis Description | Reinfection Incidence | No Reinfection Incidence | Risk Ratio | Risk Difference |
| --- | --- | --- | --- | --- |
| Primary Analysis | 0.111 | 0.082 | 1.35 (1.32, 1.39) | 0.029 (0.027, 0.031) |
| Vaccination Status |  |  |  |  |
| No Documented Vaccinations Prior to Index | 0.063 | 0.047 | 1.33 (1.23, 1.45) | 0.016 (0.011, 0.020) |
| Only Vaccinated Prior to First Infection | 0.128 | 0.088 | 1.45 (1.31, 1.60) | 0.040 (0.029, 0.050) |
| Vaccinated After First Infection Before Index | 0.072 | 0.060 | 1.19 (1.10, 1.30) | 0.012 (0.006, 0.017) |
| Vaccinated Before First Infection and Before Index | 0.154 | 0.127 | 1.21 (1.06, 1.41) | 0.027 (0.007, 0.047) |
| Negative Control Conditions |  |  |  |  |
| Papanicolaou Test | 0.002 | 0.002 | 1.17 (1.02, 1.36) | 0.000 (0.000, 0.001) |
| Non-pathological Fracture | 0.011 | 0.010 | 1.08 (1.00, 1.15) | 0.001 (0.000, 0.001) |
| Sex |  |  |  |  |
| Female | 0.116 | 0.086 | 1.34 (1.30, 1.38) | 0.029 (0.026, 0.032) |
| Male | 0.103 | 0.074 | 1.39 (1.33, 1.46) | 0.029 (0.025, 0.033) |
| Age (in years) |  |  |  |  |
| 18-25 | 0.039 | 0.028 | 1.37 (1.20, 1.57) | 0.011 (0.006, 0.015) |
| 26-35 | 0.068 | 0.049 | 1.39 (1.29, 1.50) | 0.019 (0.015, 0.023) |
| 36-50 | 0.111 | 0.077 | 1.44 (1.38, 1.52) | 0.034 (0.030, 0.039) |
| 51-65 | 0.138 | 0.101 | 1.36 (1.30, 1.42) | 0.037 (0.032, 0.042) |
| 66+ | 0.185 | 0.148 | 1.25 (1.19, 1.31) | 0.036 (0.028, 0.045) |
| Race and Ethnicity |  |  |  |  |
| American Indian or Alaska Native Non-Hispanic | 0.107 | 0.165 | 0.65 (0.35, 1.19) | -0.058 (-0.133, 0.021) |
| Asian Non-Hispanic | 0.100 | 0.088 | 1.14 (0.90, 1.41) | 0.012 (-0.009, 0.032) |
| Black or African American Non-Hispanic | 0.121 | 0.088 | 1.37 (1.28, 1.47) | 0.033 (0.026, 0.040) |
| Hispanic or Latino Any Race | 0.097 | 0.066 | 1.46 (1.32, 1.60) | 0.031 (0.023, 0.038) |
| Native Hawaiian or Other Pacific Islander Non-Hispanic | 0.176 | 0.116 | 1.52 (0.86, 2.73) | 0.060 (-0.018, 0.130) |
| Other Non-Hispanic | 0.068 | 0.048 | 1.42 (1.09, 1.98) | 0.020 (0.005, 0.039) |
| Unknown | 0.093 | 0.069 | 1.34 (1.07, 1.69) | 0.024 (0.006, 0.042) |
| White Non-Hispanic | 0.113 | 0.084 | 1.35 (1.31, 1.39) | 0.029 (0.026, 0.032) |
| Charlson Comorbidity Index |  |  |  |  |
| 0 | 0.069 | 0.049 | 1.40 (1.34, 1.45) | 0.019 (0.017, 0.022) |
| 1-2 | 0.170 | 0.123 | 1.38 (1.32, 1.44) | 0.046 (0.041, 0.053) |
| 3-4 | 0.255 | 0.202 | 1.26 (1.19, 1.36) | 0.053 (0.039, 0.068) |
| 5+ | 0.374 | 0.299 | 1.25 (1.17, 1.34) | 0.075 (0.052, 0.097) |
| Number of Visit Days in Year Before First COVID-19 Event |  |  |  |  |
| 0 | 0.049 | 0.035 | 1.40 (1.17, 1.69) | 0.014 (0.007, 0.021) |
| 1-2 | 0.044 | 0.027 | 1.60 (1.39, 1.86) | 0.017 (0.012, 0.022) |
| 3-5 | 0.047 | 0.031 | 1.50 (1.33, 1.69) | 0.016 (0.011, 0.020) |
| 6-12 | 0.065 | 0.045 | 1.45 (1.35, 1.57) | 0.020 (0.016, 0.024) |
| 13-23 | 0.093 | 0.069 | 1.35 (1.27, 1.43) | 0.024 (0.019, 0.029) |
| 24-54 | 0.148 | 0.110 | 1.34 (1.28, 1.40) | 0.037 (0.032, 0.043) |
| 55+ | 0.269 | 0.208 | 1.30 (1.25, 1.36) | 0.062 (0.053, 0.072) |
| First Infection Severity |  |  |  |  |
| No Hospitalization or Emergency Department | 0.097 | 0.072 | 1.35 (1.31, 1.39) | 0.025 (0.023, 0.028) |
| Emergency Department with No Hospitalization | 0.144 | 0.106 | 1.35 (1.28, 1.43) | 0.038 (0.031, 0.044) |
| Moderate Hospitalization | 0.253 | 0.182 | 1.39 (1.29, 1.50) | 0.071 (0.056, 0.087) |
| Severe Hospitalization with Ventilation or ECMO | 0.316 | 0.158 | 2.00 (0.78, 8.22) | 0.158 (-0.055, 0.385) |
| Quarter of First COVID-19 Event |  |  |  |  |
| 2020 Quarter 1 | 0.092 | 0.109 | 0.84 (0.49, 1.38) | -0.017 (-0.067, 0.033) |
| 2020 Quarter 2 | 0.090 | 0.066 | 1.36 (1.15, 1.60) | 0.024 (0.011, 0.036) |
| 2020 Quarter 3 | 0.089 | 0.060 | 1.48 (1.31, 1.68) | 0.029 (0.020, 0.038) |
| 2020 Quarter 4 | 0.075 | 0.054 | 1.38 (1.30, 1.46) | 0.021 (0.017, 0.024) |
| 2021 Quarter 1 | 0.088 | 0.069 | 1.27 (1.17, 1.37) | 0.019 (0.012, 0.025) |
| 2021 Quarter 2 | 0.087 | 0.065 | 1.34 (1.16, 1.56) | 0.022 (0.011, 0.034) |
| 2021 Quarter 3 | 0.106 | 0.075 | 1.42 (1.33, 1.53) | 0.032 (0.025, 0.039) |
| 2021 Quarter 4 | 0.116 | 0.085 | 1.36 (1.28, 1.46) | 0.031 (0.024, 0.037) |
| 2022 Quarter 1 | 0.147 | 0.104 | 1.41 (1.33, 1.50) | 0.043 (0.035, 0.051) |
| 2022 Quarter 2 | 0.187 | 0.151 | 1.24 (1.14, 1.35) | 0.036 (0.023, 0.051) |
| 2022 Quarter 3 | 0.211 | 0.162 | 1.30 (1.18, 1.42) | 0.049 (0.031, 0.065) |
| 2022 Quarter 4 | 0.234 | 0.185 | 1.27 (1.11, 1.48) | 0.049 (0.022, 0.080) |
| 2023 Quarter 1 | 0.301 | 0.235 | 1.28 (1.02, 1.64) | 0.065 (0.005, 0.127) |
| 2023 Quarter 2 | 0.292 | 0.324 | 0.90 (0.50, 1.84) | -0.033 (-0.218, 0.160) |
| 2023 Quarter 3 | 0.553 | 1.000 | 0.55 (0.32, 0.75) | -0.447 (-0.681, -0.248) |

We found statistically significant effects in both of our negative control conditions, though each bordered on insignificance at 95% confidence. The association between reinfections and Pap tests had a relative risk of 1.17 (95% CI, 1.02-1.36) and associations with non-pathological fractures had a relative risk of 1.08 (95% CI, 1.00-1.15). Absolute risk differences were small, reflecting the low incidence of each condition: 0.0004 (95% CI, 0.0000-0.0007) for Pap tests and 0.0008 (0.0001-0.0015) for non-pathological fractures.

The relative risk ratio of reinfections compared to non-reinfected individuals on long COVID incidence was similar across most age groups, ranging from 1.25 (95% CI, 1.19-1.31) among those 65+ years old to 1.44 (95% CI, 1.38-1.52) among those 36-50 years old. However, given differences in the cumulative incidence between these groups, we observed substantial differences in their absolute risk differences. The youngest groups have lower risk differences (18-25 years old: 0.011 [0.006-0.015]; 26-35 years old: 0.019 [0.015-0.023]) compared to older groups (36-50 years old: 0.034 [0.030-0.039]; 51-65 years old: 0.037 [0.032, 0.042]; 66+ years old: 0.036 [0.028-0.045]).

A summary of the high-confidence vaccination subanalysis cohort is shown in eTable 2. Risk ratios overlapped for most vaccination groups but were generally lower among those vaccinated post-first infection but pre-index (1.19, 1.10-1.30) and pre-first infection as well as pre-index (1.21, 1.06-1.41). Ratios were higher for those vaccinated pre-first infection but not pre-index (1.45, 1.31-1.60) or not at all (1.33, 1.23-1.45).

## Discussion

We found that SARS-CoV-2 reinfections led to a 35% (95% CI, 32-39%) higher incidence of long COVID compared to not being reinfected in an observational cohort of 424,616 individuals; 212,313 pairs of matched reinfected and non-reinfected cases. We observed a cumulative incidence of long COVID of 11.1% among reinfected individuals and 8.2% among non-reinfected individuals one year after index. Long COVID cases were expected in both cohorts— outcomes observed among non-reinfected individuals could have resulted from their first infection, an unrecorded reinfection, or model error resulting from symptoms like long COVID not caused by long COVID.

Our negative control experiments confirmed that our matching procedure, while dramatically reducing sample size, reduced utilization bias between the reinfected and non-reinfected cohorts. The relative risk of each of our negative control outcomes (Pap test procedures and non-pathological fractures), bordered on insignificance (Table 3). A key challenge was identifying sufficiently prevalent negative control conditions with no suspected link to COVID-19 or long COVID, which have been linked to cardiorespiratory, cognitive, gastrointestinal, musculoskeletal, autoimmune, and other symptoms.^31,32^ We relied on physicians and patient advocates to validate our selections. However, a recent study found that long COVID may be associated with some types of fractures.^33^

Our primary result, that reinfections lead to a greater risk of long COVID, were similar to those reported by Bowe, Xie, and Al-Aly in 2022, the only large, published study we know of quantifying the effect of reinfections on long COVID.^7^ However, our study differs from theirs in several key respects. First, their study uses the United States Department of Veterans Affairs’ national healthcare database, which is 90% male, as well as disproportionately white and older. Our cohort is more diverse in age, sex, race and ethnicity. Second, Bowe et al. included reinfections from June 1, 2020 to June 25, 2022. Our study used more recent data and was limited to reinfections in the Omicron era, between November 1, 2021 and June 1, 2024. Third, they used an extensive covariate list to construct logistic regressions for propensity score calculation, then used the propensity scores for inverse probability of treatment weighting. We elected to use coarsened exact matching to capture all possible interaction effects in a small set of key confounders. Our use of one-to-one matching allowed us to assign index dates to non-reinfected cases aligned with matched controls, as well as to censor both cases when either is lost to follow-up. We also did not require that non-reinfected participants have no future reinfections, preventing bias in cohort allocation. Fourth, our definition of long COVID was based on a computable phenotype, which used a machine learning model to identify individuals whose symptoms closely align with individuals clinically diagnosed with long COVID, rather than the incidence of any symptom that has been linked to long COVID. Our results strengthen the finding that reinfections increase the risk of long COVID.

Risk ratios were consistent across most strata (Table 3); individuals in strata with higher overall incidence therefore experienced larger differences in absolute risk. An exception was site of care, which included wide ranges of risk ratios driven by high cardinality, small sample sizes, and differences in geography and provider behavior (eTable 1). We did not interpret differences in site of care, as it is included as a matching criterion and sites are anonymized to preserve participant privacy. Variation across sites suggests a potential for site-specific factors that warrant further investigation.

In the vaccination subanalysis, we stratified by vaccination status into four groups: unvaccinated prior to index date, vaccinated only prior to first infection, vaccinated only between first infection and index date, and vaccinated both prior to first infection and between first infection and index date. Risk ratio confidence intervals for all groups overlapped, except for between those vaccinated between first infection and index and those vaccinated only prior to their first infection. Those vaccinated between first infection and index faced lower risks of long COVID from reinfection than those only vaccinated prior to their first infection, suggesting that more recent vaccinations may lower the risk of long COVID following reinfections. However, this effect was not consistent across all groups. Vaccinated individuals tended to be older with more comorbidities and utilization (eTable 2), resulting in higher overall incidences of long COVID. More research is needed to discern how vaccination modulates the likelihood of a reinfection to lead to long COVID.

### Limitations

We are limited by our use of EHRs to capture matching criteria, reinfection status, and features informing our long COVID computable phenotype. EHRs may not be broadly representative and have well-documented data quality issues that can lead to bias and misinterpretations.^34,35^ Individuals receiving care outside of the site providing their data had incomplete records. We could not include reinfections that were not observed (including home tests) or did not result in a documented positive lab test. It is possible that some of our control cases experienced reinfections, potentially dampening our observed effect of reinfection.

Like all causal studies using observational data, we were limited to including confounders available in our data. Insofar as unmeasured confounders operate independently from our included matching criteria, they have the potential to bias our results. A prominent latent confounder is the propensity and ability to seek care, which affects both the likelihood of COVID-19 reinfection and long COVID symptoms being documented in the EHR. However, our inclusion of pre-COVID-19 CCI and counts of visit days in the two years prior to first COVID-19 event minimize this effect.

While we believe our use of a computable phenotype for long COVID is a strength, as outlined in the methods, it is likely that some participants were misclassified. Since we did not account for this uncertainty in our bootstrap sampling, our confidence intervals may be underestimated. The computable phenotype also does not assess severity, so we are unable to conclude if long COVID after reinfection differs in severity from first infection.

We limited our study to comparing the effect of first reinfections on risk of long COVID. We did not measure the impact of subsequent reinfections due to having a limited sample of observed cases. We cannot guarantee that individuals did not experience reinfections before their first recorded reinfection. We also only include adults; how reinfections change long COVID incidence in children should be addressed in future research.

In conclusion, we found that reinfections lead to a roughly 35% (95% CI, 32-39%) increase in the risk of long COVID as compared to not having a documented reinfection. The relative effect was consistent across most strata of most of our matching criteria, including sex and timing and severity of first infection. There were differences in relative risk across different age and CCI strata. In a vaccination subanalysis, we found that those vaccinated between first infection and index date experienced a smaller increase in the relative risk of long COVID resulting from reinfection as compared to those vaccinated only prior to their first infection.

## Supporting information

Statistical Analysis Plan

## Data Availability

All data used and produced in the present work are available online to registered users of the National Clinical Cohort Collaborative (N3C). More information on how to register and obtain access can be found at https://covid.cd2h.org/for-researchers

## Acknowledgements

This study is part of the NIH Researching COVID to Enhance Recovery (RECOVER) Initiative, which seeks to understand, treat, and prevent the post-acute sequelae of SARS-CoV-2 infection. For more information on RECOVER, visit https://recovercovid.org/. This research was funded by the National Institutes of Health (NIH) Agreement OTA OT2HL161847 as part of the Researching COVID to Enhance Recovery (RECOVER) research program.

Authors have no conflicts of interest to disclose. Authorship was determined according to ICMJE recommendations.

The analyses described in this publication were conducted with data or tools accessed through the NCATS N3C Data Enclave https://covid.cd2h.org and N3C Attribution & Publication Policy v 1.2-2020-08-25b supported by NCATS Contract No. 75N95023D00001, Axle Informatics Subcontract: NCATS-P00438-B, and by the RECOVER Initiative (OT2HL161847–01). This research was possible because of the patients whose information is included within the data and the organizations (https://ncats.nih.gov/n3c/resources/data-contribution/data-transfer-agreement-signatories) and scientists who have contributed to the on-going development of this community resource [https://doi.org/10.1093/jamia/ocaa196].

The N3C Publication committee confirmed that this manuscript msid:2453.184 is in accordance with N3C data use and attribution policies; however, this content is solely the responsibility of the authors and does not necessarily represent the official views of the National Institutes of Health or the N3C program.

## IRB

The N3C data transfer to NCATS is performed under a Johns Hopkins University Reliance Protocol # IRB00249128 or individual site agreements with NIH. The N3C Data Enclave is managed under the authority of the NIH; information can be found at https://ncats.nih.gov/n3c/resources.

N3C consortial contributors: Christopher G. Chute

RECOVER consortial contributors: Yu Chen

We gratefully acknowledge the following core contributors to N3C:

Adam B. Wilcox, Adam M. Lee, Alexis Graves, Alfred (Jerrod) Anzalone, Amin Manna, Amit Saha, Amy Olex, Andrea Zhou, Andrew E. Williams, Andrew M. Southerland, Andrew T. Girvin, Anita Walden, Anjali Sharathkumar, Benjamin Amor, Benjamin Bates, Brian Hendricks, Brijesh Patel, G. Caleb Alexander, Carolyn T. Bramante, Cavin Ward-Caviness, Charisse Madlock-Brown, Christine Suver, Christopher G. Chute, Christopher Dillon, Chunlei Wu, Clare Schmitt, Cliff Takemoto, Dan Housman, Davera Gabriel, David A. Eichmann, Diego Mazzotti, Donald E. Brown, Eilis Boudreau, Elaine L. Hill, Emily Carlson Marti, Emily R. Pfaff, Evan French, Farrukh M Koraishy, Federico Mariona, Fred Prior, George Sokos, Greg Martin, Harold P. Lehmann,

Heidi Spratt, Hemalkumar B. Mehta, J.W. Awori Hayanga, Jami Pincavitch, Jaylyn Clark, Jeremy Richard Harper, Jessica Yasmine Islam, Jin Ge, Joel Gagnier, Johanna J. Loomba, John B. Buse, Jomol Mathew, Joni L. Rutter, Julie A. McMurry, Justin Guinney, Justin Starren, Karen Crowley, Katie Rebecca Bradwell, Kellie M. Walters, Ken Wilkins, Kenneth R. Gersing, Kenrick Cato, Kimberly Murray, Kristin Kostka, Lavance Northington, Lee Pyles, Lesley Cottrell, Lili M. Portilla, Mariam Deacy, Mark M. Bissell, Marshall Clark, Mary Emmett, Matvey B. Palchuk, Melissa A. Haendel, Meredith Adams, Meredith Temple-O’Connor, Michael G. Kurilla, Michele Morris, Nasia Safdar, Nicole Garbarini, Noha Sharafeldin, Ofer Sadan, Patricia A. Francis, Penny Wung Burgoon, Philip R.O. Payne, Randeep Jawa, Rebecca Erwin-Cohen, Rena C. Patel, Richard A. Moffitt, Richard L. Zhu, Rishikesan Kamaleswaran, Robert Hurley, Robert T. Miller, Saiju Pyarajan, Sam G. Michael, Samuel Bozzette, Sandeep K. Mallipattu, Satyanarayana Vedula, Scott Chapman, Shawn T. O’Neil, Soko Setoguchi, Stephanie S. Hong, Steven G. Johnson, Tellen D. Bennett, Tiffany J. Callahan, Umit Topaloglu, Valery Gordon, Vignesh Subbian, Warren A. Kibbe, Wenndy Hernandez, Will Beasley, Will Cooper, William Hillegass, Xiaohan Tanner Zhang. Details of contributions available at covid.cd2h.org/core-contributors

Data Partners with Released Data

The following institutions whose data is released or pending:

### Available

Advocate Health Care Network — UL1TR002389: The Institute for Translational Medicine (ITM) • Aurora Health Care Inc — UL1TR002373: Wisconsin Network For Health Research • Boston University Medical Campus — UL1TR001430: Boston University Clinical and Translational Science Institute • Brown University — U54GM115677: Advance Clinical Translational Research (Advance-CTR) • Carilion Clinic — UL1TR003015: iTHRIV Integrated Translational health Research Institute of Virginia • Case Western Reserve University — UL1TR002548: The Clinical & Translational Science Collaborative of Cleveland (CTSC) • Charleston Area Medical Center — U54GM104942: West Virginia Clinical and Translational Science Institute (WVCTSI) • Children’s Hospital Colorado — UL1TR002535: Colorado Clinical and Translational Sciences Institute • Columbia University Irving Medical Center — UL1TR001873: Irving Institute for Clinical and Translational Research • Dartmouth College — None (Voluntary) Duke University — UL1TR002553: Duke Clinical and Translational Science Institute • George Washington Children’s Research Institute — UL1TR001876: Clinical and Translational Science Institute at Children’s National (CTSA-CN) • George Washington University — UL1TR001876: Clinical and Translational Science Institute at Children’s National (CTSA-CN) • Harvard Medical School — UL1TR002541: Harvard Catalyst • Indiana University School of Medicine — UL1TR002529: Indiana Clinical and Translational Science Institute • Johns Hopkins University — UL1TR003098: Johns Hopkins Institute for Clinical and Translational Research • Louisiana Public Health Institute — None (Voluntary) • Loyola Medicine — Loyola University Medical Center • Loyola University Medical Center — UL1TR002389: The Institute for Translational Medicine (ITM) • Maine Medical Center — U54GM115516: Northern New England Clinical & Translational Research (NNE-CTR) Network • Mary Hitchcock Memorial Hospital & Dartmouth Hitchcock Clinic — None (Voluntary) • Massachusetts General Brigham — UL1TR002541: Harvard Catalyst • Mayo Clinic RochesterUL1TR002377: Mayo Clinic Center for Clinical and Translational Science (CCaTS) • Medical University of South Carolina — UL1TR001450: South Carolina Clinical & Translational Research Institute (SCTR) • MITRE Corporation — None (Voluntary) • Montefiore Medical Center — UL1TR002556: Institute for Clinical and Translational Research at Einstein and Montefiore • Nemours — U54GM104941: Delaware CTR ACCEL Program • NorthShore University HealthSystem — UL1TR002389: The Institute for Translational Medicine (ITM) • Northwestern University at Chicago — UL1TR001422: Northwestern University Clinical and Translational Science Institute (NUCATS) • OCHIN — INV-018455: Bill and Melinda Gates Foundation grant to Sage Bionetworks • Oregon Health & Science University — UL1TR002369: Oregon Clinical and Translational Research Institute • Penn State Health Milton S. Hershey Medical Center — UL1TR002014: Penn State Clinical and Translational Science Institute • Rush University Medical Center — UL1TR002389: The Institute for Translational Medicine (ITM) Rutgers, The State University of New Jersey — UL1TR003017: New Jersey Alliance for Clinical and Translational Science • Stony Brook University — U24TR002306 • The Alliance at the University of Puerto Rico, Medical Sciences Campus — U54GM133807: Hispanic Alliance for Clinical and Translational Research (The Alliance) • The Ohio State University — UL1TR002733: Center for Clinical and Translational Science • The State University of New York at Buffalo — UL1TR001412: Clinical and Translational Science Institute • The University of Chicago — UL1TR002389: The Institute for Translational Medicine (ITM) • The University of Iowa — UL1TR002537: Institute for Clinical and Translational Science • The University of Miami Leonard M. Miller School of Medicine — UL1TR002736: University of Miami Clinical and Translational Science Institute • The University of Michigan at Ann Arbor — UL1TR002240: Michigan Institute for Clinical and Health Research • The University of Texas Health Science Center at Houston — UL1TR003167: Center for Clinical and Translational Sciences (CCTS) • The University of Texas Medical Branch at Galveston — UL1TR001439: The Institute for Translational Sciences • The University of Utah — UL1TR002538: Uhealth Center for Clinical and Translational Science • Tufts Medical Center — UL1TR002544: Tufts Clinical and Translational Science Institute • Tulane University — UL1TR003096: Center for Clinical and Translational Science • The Queens Medical Center — None (Voluntary) • University Medical Center New Orleans — U54GM104940: Louisiana Clinical and Translational Science (LA CaTS) Center • University of Alabama at Birmingham — UL1TR003096: Center for Clinical and Translational Science • University of Arkansas for Medical Sciences — UL1TR003107: UAMS Translational Research Institute • University of Cincinnati — UL1TR001425: Center for Clinical and Translational Science and Training • University of Colorado Denver, Anschutz Medical Campus — UL1TR002535: Colorado Clinical and Translational Sciences Institute • University of Illinois at Chicago — UL1TR002003: UIC Center for Clinical and Translational Science • University of Kansas Medical Center — UL1TR002366: Frontiers: University of Kansas Clinical and Translational Science Institute • University of Kentucky — UL1TR001998: UK Center for Clinical and Translational Science • University of Massachusetts Medical School Worcester — UL1TR001453: The UMass Center for Clinical and Translational Science (UMCCTS) • University Medical Center of Southern Nevada — None (voluntary) • University of Minnesota — UL1TR002494: Clinical and Translational Science Institute • University of Mississippi Medical Center — U54GM115428: Mississippi Center for Clinical and Translational Research (CCTR) • University of Nebraska Medical Center — U54GM115458: Great Plains IDeA-Clinical & Translational Research • University of North Carolina at Chapel Hill — UL1TR002489: North Carolina Translational and Clinical Science Institute • University of Oklahoma Health Sciences Center — U54GM104938: Oklahoma Clinical and Translational Science Institute (OCTSI) • University of Pittsburgh — UL1TR001857: The Clinical and Translational Science Institute (CTSI) • University of Pennsylvania — UL1TR001878: Institute for Translational Medicine and Therapeutics • University of Rochester — UL1TR002001: UR Clinical & Translational Science Institute • University of Southern California — UL1TR001855: The Southern California Clinical and Translational Science Institute (SC CTSI) • University of Vermont — U54GM115516: Northern New England Clinical & Translational Research (NNE-CTR) Network • University of Virginia — UL1TR003015: iTHRIV Integrated Translational health Research Institute of Virginia University of Washington — UL1TR002319: Institute of Translational Health Sciences • University of Wisconsin-Madison — UL1TR002373: UW Institute for Clinical and Translational Research • Vanderbilt University Medical Center — UL1TR002243: Vanderbilt Institute for Clinical and Translational Research • Virginia Commonwealth University — UL1TR002649: C. Kenneth and Dianne Wright Center for Clinical and Translational Research • Wake Forest University Health Sciences — UL1TR001420: Wake Forest Clinical and Translational Science Institute • Washington University in St. Louis — UL1TR002345: Institute of Clinical and Translational Sciences • Weill Medical College of Cornell University — UL1TR002384: Weill Cornell Medicine Clinical and Translational Science Center • West Virginia University — U54GM104942: West Virginia Clinical and Translational Science Institute (WVCTSI) Submitted: Icahn School of Medicine at Mount Sinai — UL1TR001433: ConduITS Institute for Translational Sciences • The University of Texas Health Science Center at Tyler — UL1TR003167: Center for Clinical and Translational Sciences (CCTS) • University of California, Davis — UL1TR001860: UCDavis Health Clinical and Translational Science Center • University of California, Irvine — UL1TR001414: The UC Irvine Institute for Clinical and Translational Science (ICTS) • University of California, Los Angeles — UL1TR001881: UCLA Clinical Translational Science Institute • University of California, San Diego — UL1TR001442: Altman Clinical and Translational Research Institute • University of California, San Francisco — UL1TR001872: UCSF Clinical and Translational Science Institute NYU Langone Health Clinical Science Core, Data Resource Core, and PASC Biorepository Core — OTA-21-015A: Post-Acute Sequelae of SARS-CoV-2 Infection Initiative (RECOVER)

### Pending

Arkansas Children’s Hospital — UL1TR003107: UAMS Translational Research Institute • Baylor College of Medicine — None (Voluntary) • Children’s Hospital of PhiladelphiaUL1TR001878: Institute for Translational Medicine and Therapeutics • Cincinnati Children’s Hospital Medical Center — UL1TR001425: Center for Clinical and Translational Science and Training • Emory University — UL1TR002378: Georgia Clinical and Translational Science Alliance • HonorHealth — None (Voluntary) • Loyola University Chicago — UL1TR002389: The Institute for Translational Medicine (ITM) • Medical College of Wisconsin — UL1TR001436: Clinical and Translational Science Institute of Southeast Wisconsin • MedStar Health Research Institute — None (Voluntary) • Georgetown University — UL1TR001409: The Georgetown-Howard Universities Center for Clinical and Translational Science (GHUCCTS) • MetroHealth — None (Voluntary) • Montana State University — U54GM115371: American Indian/Alaska Native CTR • NYU Langone Medical Center — UL1TR001445: Langone Health’s Clinical and Translational Science Institute • Ochsner Medical Center — U54GM104940: Louisiana Clinical and Translational Science (LA CaTS) Center • Regenstrief Institute — UL1TR002529: Indiana Clinical and Translational Science Institute • Sanford Research — None (Voluntary) • Stanford University — UL1TR003142: Spectrum: The Stanford Center for Clinical and Translational Research and Education • The Rockefeller University — UL1TR001866: Center for Clinical and Translational Science • The Scripps Research Institute — UL1TR002550: Scripps Research Translational Institute • University of Florida — UL1TR001427: UF Clinical and Translational Science Institute • University of New Mexico Health Sciences Center — UL1TR001449: University of New Mexico Clinical and Translational Science Center • University of Texas Health Science Center at San Antonio — UL1TR002645: Institute for Integration of Medicine and Science • Yale New Haven Hospital — UL1TR001863: Yale Center for Clinical Investigation

