## Supplementary material for "Incidence of Long COVID Following Reinfection with COVID-19": Statistical Analysis Plan

### Data

All analysis was conducted using data from the National Cohort Clinical Collaborative.[1] Information on how to access the data and code for replication or original analyses can be found at https://covid.cd2h.org/for-researchers/.

### Final Analysis Plan

| Concept | Definition |
|---|---|
| **Step 1: Identify study population**<br>Identify patients meeting inclusion and exclusion criteria. | |
| **Study period** | 3/1/2020 – 6/1/2024 |
| **Evidence of COVID-19** | Positive SARS-CoV-2 PCR or antigen test, prescription for nirmatrelvir-ritonavir (Paxlovid), or ≥1 U07.1 ICD-10 code |
| **Enrollment date** | First evidence of COVID-19 between 3/1/2020 and 12/4/2023. Cutoff of 12/4/2023 was chosen to allow for 180 days of follow-up after enrollment, and most sites of care included data through 6/1/2024. |
| **Inclusion criteria** | • Has a valid enrollment date<br>• Between 18 and 99 years old at enrollment date |
| **Exclusion criteria** | • Patients with missing or unknown sex<br>• Long COVID outcome prior to enrollment date<br>• See additional exclusion in Step 3 |
| **Step 2: Matching**<br>Assign individuals into matched pairs. | |
| **Matching Criteria** | Calculated at enrollment date, with continuous features discretized into the provided ranges.<br>• Sex: male or female<br>• Age: 18-25, 26-35, 36-50, 51-65, 66+<br>• Race/ethnicity (all non-Hispanic unless indicated otherwise): American Indian or Alaskan native, Asian, Black, Hispanic any race, native Hawaiian or other pacific islander, other, unknown, White<br>• Charlson Comorbidity Index: 0, 1-2, 3-4, 5+<br>• Number of visit days in the year prior to enrollment date: 0, 1-2, 3-5, 6-12, 13-23, 24-54, 55+<br>• Severity of first COVID-19 infection: no hospitalization or emergency department (ED), ED with no hospitalization, hospitalization without ventilation or extracorporeal membrane oxygenation (ECMO), hospitalization with ventilation or ECMO<br>• Quarter of first COVID-19 infection (e.g., 2020 Q1, 2020 Q2, etc.)<br>• Site of care |

| | |
|---|---|
| **Matching Procedure** | Group individuals according to alignment in all matching criteria. Within each group, randomly assign one unexposed individual to each reinfected individual to form matched pairs. Make assignments one-at-a-time, in chronological order of dates of reinfection among reinfected individuals. At the time of each pairing, all unpaired individuals are eligible to be selected as a matched unexposed case. Individuals with later reinfections are therefore able to serve as unexposed cases for earlier reinfections. Thus, unexposed cases are not selected based on a future lack of reinfections. Drop individuals not used as a reinfected nor unexposed case. |
| **Index date** | For reinfected individuals: First positive SARS-CoV-2 PCR or antigen test occurring at least 60 days after enrollment.<br>For unexposed individuals: Uses the index date of their matched reinfected individual |
| **Step 3: Assess outcomes**<br>Assign outcome status. | |
| **Outcome** | Use the updated computable phenotype developed by Pfaff et al.[2] Scores between 0 and 1 are generated for 100-day evaluation windows in 30-day increments. Higher scores indicate closer alignment with health records of diagnosed long COVID cases. A score above 0.9 in any window indicates sufficient evidence of long COVID, with the onset date assigned as the beginning of the evaluation window of the highest-scoring period. |
| **Exclusion criterion** | Exclude any reinfected or unexposed individual that has the outcome prior to their index date, along with their matched individual. |
| **Step 4: Statistical Analysis**<br>Quantify the effect of exposure. | |
| **Censoring Events** | Censor both individuals within a matched pair at the earliest of any censoring event:<br>• A new reinfection<br>• Last recorded visit, prescription, or diagnosis<br>• Death<br>• End of the study period (6/1/2024) |
| **Cumulative Incidence** | Use Aalen-Johansen estimator to calculate a cumulative incidence of the outcome among reinfected individuals and among exposure individuals after 365 days. |
| **Effect Estimates** | • Risk ratio: cumulative incidence among reinfected individuals divided by the cumulative incidence among unexposed individuals<br>• Risk difference: cumulative incidence among reinfected individuals subtracted by the cumulative incidence among unexposed individuals |
| **Negative Control Analysis**<br>Validate the significance of effects with negative control outcomes. | |
| **Negative Control Outcomes** | Administration of a Papanicolaou screening test: presence of the standard OMOP code 2720580, corresponding to the HCPCS code Q0091.<br>Evidence of a non-pathological fracture: any fracture diagnosis that is not indicate as a pathological fracture. |
| **Analysis** | Repeat Steps 3-4 using each of the negative control outcomes in place of the long COVID outcome. |

| **Vaccine Subanalysis**<br>Estimate whether COVID-19 vaccination modulates the effect of reinfections on the long COVID outcome. | |
|---|---|
| **High-confidence vaccination sites** | Calculate the proportion of people with at least one record of COVID-19 vaccination at a site of care. Compare the site vaccination rate to age and county-level vaccination rates reported by the U.S. Centers for Disease Control (this data is no longer available).[3] Retain sites where the site vaccination rate is at least 2/3 of the CDC-reported rate. |
| **Vaccination Status** | Vaccination status falls into one of four groups:<br>• Pre-enrollment and pre-index vaccination: at least one recorded vaccination both prior to the enrollment date and between the enrollment and index dates.<br>• Pre-enrollment without pre-index vaccination: at least one recorded vaccination prior to the enrollment date with no additional vaccinations between the enrollment and index dates.<br>• Pre-index without pre-enrollment vaccination: no recorded vaccinations prior to the enrollment date with at least one vaccination between the enrollment and index dates.<br>• No vaccinations: No recorded vaccinations prior to the index date. |
| **Matching** | Matching is performed as in the primary analysis with the following changes:<br>• Pre-enrollment vaccination status (at least one vaccination or no recorded vaccinations) is added to the matching criteria.<br>• After matching, remove any pairs with misaligned vaccination status between enrollment and index. |

### Original Analysis Plan

This study evolved from a short-term ad hoc request from the RECOVER Initiative to a full-scale study. Below are specifications for the original analysis. The original analysis included collaborators using additional data sources, who did not participate in the final study.

#### Version History

| Version | Date | Notes |
|---|---|---|
| 1.0 | 5/30/2023 | Initial draft created |
| 1.1 | 6/6/2023 | Draft ready for review |
| 1.2 | 6/22/2023 | Modified specifications to remove timing component from subphenotype assessment, and switch long COVID outcome to computable phenotype definition, added additional items to Table 1 |
| 1.3 | 6/26/2023 | Updated open items and Figure 4 description |
| 1.4 | 7/7/2023 | Replaced Figure 3 (infections over time) with Sankey diagram, modified Figure 2 to replace prior table looking at time between reinfections. |

#### Description

The objective of this analysis is to examine the association between the number, severity, and variant era of SARS-CoV-2 infections and the risk of PASC. Specifically, it will focus on:

- Descriptive output (e.g, number of reinfections, time between reinfections), overall and by PASC status
- The relationship between reinfections and PASC subphenotypes

### Procedure

| Concept | Definition |
|---|---|
| **Step 1: Identify study population**<br>Identify patients meeting inclusion and exclusion criteria as described below. | |
| **Study period** | 3/1/2020 – 4/30/2023 |
| **COVID-19 positive** | Positive SARS-CoV-2 PCR or antigen test, U09.9 code, prescription for Paxlovid or remdesivir, or ≥1 U07.1 code |
| **Index event** | <u>For patients without a U09.9 code</u>: The earliest evidence of COVID-19 positivity within the study period<br><u>For patients with a U09.9 code</u>: The earliest other evidence of COVID-19 positivity within the study period; or, if no other evidence is available, impute index event as 59 days prior to the U09.9 code |
| **Inclusion criteria** | • COVID-19 index event prior to 10/31/2022<br>• One or more visits in the two years prior to index<br>• At least one visit >60 days after the index event |
| **Exclusion criteria** | • Patients with missing or unknown sex<br>• N3C and PCORnet: <21 years old at time of index<br>• PEDSnet: ≥21 years old at time of index |
| **Step 3: Assess outcomes**<br>Query specific definitions are below. | |
| **Primary infection** | COVID index event |
| **First reinfection** | Positive SARS-CoV-2 PCR or antigen test, or Paxlovid or remdesivir order, that occurred 60 or more days after a COVID-19 infection index date. The date of the test or medication order is considered the first COVID-19 reinfection index date. |
| **Subsequent reinfection** | A new positive SARS-CoV-2 PCR or antigen test, or evidence of Paxlovid or remdesivir, occurring 60 or more days after each reinfection index date |
| **PASC** | Apply each Cohorts' non-harmonized PASC definition within the study period. First qualifying evidence of PASC should be considered the PASC start date.<br><br>If Cohorts can run PASC definitions with blackout periods around reinfections, please implement as a sensitivity analysis with Table 1 and Figure 2. |
| **Step 4: Produce Table 1**<br>Query specific definitions are below. | |
| **COVID severity** | • <u>Hospitalized</u>: Patient was hospitalized within 1 day before to 16 days after the COVID index event, where the diagnoses include evidence of COVID-19 infection |

| | |
|---|---|
| | • Hospitalized with ventilation: Hospitalized with evidence of ICU level care, invasive ventilation/ECMO, or vasopressor/inotropic support<br>• Not hospitalized: No evidence of hospitalization at index event |
| **COVID treatment** | Evidence of Paxlovid or remdesivir at any time in the study period |
| **Step 5: Generate output**<br>See below for proposed output. | |

**Standard Table 1. Attrition Table (directional)**

| Criteria | # Patients |
|---|---|
| Patients with available EHR data in Network | |
| COVID-19 index event prior to 4/30/2022 | |
| One or more visits in the two years prior to index event | |
| At least one visit >60 days after the index event | |
| Did not have missing/unknown sex | |
| Final count | |

**Table 1. Characteristics of COVID+ patients by reinfection status**

| Characteristic | All patients # (Col%;Row%) | Without documented reinfection # (Col%;Row%) | With reinfection # (Col%;Row%) |
|---|---|---|---|
| **N** | | | |
| **Mean age (± SD)** | | | |
| **Age groups** | | | |
| <1 | | | |
| 1-4 | | | |
| 5-9 | | | |
| 10-15 | | | |
| 16-20 | | | |
| 21-45 | | | |
| 46-65 | | | |
| 66+ | | | |
| **Sex** | | | |
| Male | | | |
| Female | | | |
| **Race/ethnicity** | | | |
| Non-Hispanic Asian | | | |
| Non-Hispanic Black | | | |
| Non-Hispanic White | | | |
| Non-Hispanic Other | | | |
| Hispanic | | | |
| Missing/Unknown | | | |
| **Variant era of index infection** | | | |
| Ancestral (3/1/20 – 9/30/20) | | | |

| | | |
|---|---|---|
| Alpha (10/1/20 – 5/30/21) | | |
| Delta (6/1/21 – 11/30/21) | | |
| Omicron (12/1/21 – Present) | | |
| **Number of reinfections** | | |
| None | | |
| 1 | | -- |
| 2 | | -- |
| 3+ | | -- |
| **COVID Severity** | | |
| Never hospitalized | | |
| Hospitalized without ventilation or IMV/ECMO/vasopressor use | | |
| Hospitalized with ventilation or IMV/ECMO/vasopressor use | | |
| **COVID Treatment** | | |
| Paxlovid | | |
| Remdesivir | | |
| **Developed PASC** | | |
| **Number of PASC subphenotypes** | | |
| 1 | | |
| 2 | | |
| 3+ | | |
| **For those with PASC: # documented reinfections prior to PASC diagnosis** | | |
| None | | |
| 1 | | |
| 2 | | |
| 3+ | | |

**Figure 1**. **Number of infections**

Create three stacked bar charts looking at documented infections among the study population, stratifying by:

A. Race/ethnicity
B. Age, and
C. Sex

*Formatting details:*

- *X-axis: Number of infections*
- *Y-axis: Percentage of patients*
- *Different colors to represent race/ethnicity, age, and sex; see example from prior query below*

**Figure 2**. **Infections by variant era**

Graph the frequency of primary infections, first reinfections, second reinfections by variant era, for:

- Patients without PASC
- Patients with PASC

**Figure 3. Reinfections over time**

Proposing one of the following two options:

**Option 1:**

At each time point (representing 60-day periods from the index event), plot the % of COVID patients in each state. States include: COVID infection, COVID infection with hospitalization, PASC onset, deceased, and none in the past 60 days.

Notes:

- At time zero, all patients have infection or infection with hospitalization
- If a patient is deceased, they stay in that category for the duration
- If a patient develops PASC, they can still change states (e.g., to deceased or infected) in future time periods
- If a patient had two states in the same period, note the more recent
- If a patient has no recent infections, death, or PASC onset, classify them as "none"

**Option 2:**

Sankey diagram using chronological order, but without set timeframes noted in the diagram.

Nodes should be the following:

1. Hospitalized or non-hospitalized initial infection
2. Patient had reinfection, developed PASC, or died
3. Continue until patients have died, reached the third reinfection, or have no additional data/status changes

**Figure 4.** PASC patient subphenotype profiles by reinfection status

Produce stacked bar charts capturing the percentage of PASC patients with each subphenotype, stratifying by patients having reinfection vs. no documented reinfection, for:

1. Patients having one subphenotype
2. Patients having multiple subphenotypes

### Changes and Rationale

Many changes were made to this exploratory analysis to makes its results more clinically meaningful. A summary of those changes and their rationale are presented below.

- We introduce matching for causal interpretations. The original analysis explored only associative relationships. By matching on critical criteria, we can infer a causal relationship between reinfections and long COVID incidence. This shift also required distinguishing matching criteria from the outcome of interest, resulting in a focus on long COVID as the only outcome.

- We use cumulative incidence instead of odds ratios as our effect to account for differences in exposure times for long COVID. Otherwise, traits associated with earlier reinfections may appear related to long COVID due to having more time to develop it.
- We limit to reinfections in the Omicron era to maximize relevance to current reinfections.
- We remove the ICD-10 diagnosis code for long COVID, U09.9, as an indication for COVID-19. We require at least 60 days to pass for a reinfection to occur; imputing a COVID-19 enrollment date 59 days prior to long COVID diagnosis, as described in the original protocol, does not provide adequate time for long COVID to develop after a subsequent reinfection index date.
- We remove long COVID subphenotypes as an outcome to simplify the analysis and results.
