## Supplement for "Incidence of Long COVID Following Reinfection with COVID-19"

**Outcome definition.** Our outcome is a computable phenotype based on a machine learning model.[1,2] The model labeled individuals with a likelihood of long COVID in overlapping 100-day windows according to the similarity of their recorded conditions to those with multiple clinical diagnoses of long COVID. Individuals with a score over 0.9 in any window were labeled as having long COVID. A cutoff of 0.9 maximized the Youden Index (the true positive rate minus the false positive rate) on a held-out validation set.[2] If multiple windows were above 0.9, the highest score (having the strongest evidence) was used as the outcome onset date. We did not include clinical diagnoses of long COVID (a U09.9 ICD-10 diagnosis code) as an outcome because we found high rates of U09.9 usage contemporaneous with reinfections. This co-occurrence of U09.9 and positive COVID-19 tests suggested that U09.9 may often be conflated in the context of reinfections. It is also common to use incidence of one or more symptoms linked to long COVID as a long COVID outcome, prioritizing sensitivity over specificity. We elect to use a high confidence computable phenotype as a complement to these less specific methods.

**Discussion of methods for causality.** We used coarsened exact matching to establish exchangeability between our reinfected and non-reinfected cohorts. Other methods, such as propensity score matching or inverse probability of treatment weighting, are more common, in part because of the large reduction in sample resulting from exact matching. However, despite a large reduction, our sample of 424,616 individuals remains large enough to observe a clear effect of reinfections. Both propensity score matching and inverse probability of treatment weighting reduce overall similarity to a single dimension in the form of a probability of receiving a treatment (or in our case, being reinfected). Our approach, while reducing sample size, controls for all possible interactions in critical confounders: age, sex, race, Charlson comorbidity index, healthcare utilization, timing and severity of first COVID-19 infection, and site of care.

eTable 1. Cumulative incidences, risk ratio, and risk difference stratified by site of care. Nine sites are masked due to having small counts of outcomes.

| Analysis Description | Reinfection Incidence | Control Incidence | Risk Ratio | Risk Difference |
|---|---|---|---|---|
| Site of Care | | | | |
| A | * | * | * | * |
| B | 0.174 | 0.114 | 1.52 (1.26, 1.94) | 0.060 (0.032, 0.091) |
| C | 0.129 | 0.091 | 1.41 (1.12, 1.77) | 0.037 (0.012, 0.063) |
| D | 0.126 | 0.153 | 0.83 (0.49, 1.51) | -0.026 (-0.098, 0.053) |
| E | 0.061 | 0.047 | 1.31 (1.24, 1.37) | 0.014 (0.012, 0.017) |
| F | 0.189 | 0.145 | 1.30 (1.16, 1.43) | 0.043 (0.026, 0.060) |
| G | * | * | * | * |
| H | 0.111 | 0.091 | 1.22 (0.96, 1.55) | 0.020 (-0.004, 0.045) |
| I | 0.163 | 0.124 | 1.32 (1.06, 1.69) | 0.039 (0.008, 0.073) |
| J | 0.223 | 0.173 | 1.29 (1.07, 1.65) | 0.050 (0.014, 0.095) |

| | | | | |
|---|---|---|---|---|
| K | 0.170 | 0.125 | 1.36 (1.22, 1.54) | 0.045 (0.028, 0.063) |
| L | 0.117 | 0.093 | 1.26 (0.96, 1.72) | 0.025 (-0.004, 0.057) |
| M | 0.169 | 0.120 | 1.41 (1.18, 1.67) | 0.049 (0.024, 0.073) |
| N | 0.197 | 0.125 | 1.58 (1.17, 2.24) | 0.072 (0.026, 0.121) |
| O | 0.126 | 0.080 | 1.57 (1.35, 1.87) | 0.046 (0.030, 0.062) |
| P | 0.130 | 0.082 | 1.59 (1.00, 2.49) | 0.048 (0.000, 0.093) |
| Q | 0.166 | 0.072 | 2.30 (1.36, 4.16) | 0.094 (0.032, 0.149) |
| R | 0.043 | 0.039 | 1.10 (0.64, 1.88) | 0.004 (-0.017, 0.026) |
| S | 0.146 | 0.111 | 1.31 (1.14, 1.50) | 0.034 (0.017, 0.052) |
| T | 0.089 | 0.056 | 1.60 (1.04, 2.47) | 0.033 (0.003, 0.062) |
| U | 0.258 | 0.146 | 1.76 (1.24, 2.53) | 0.112 (0.044, 0.179) |
| V | * | * | * | * |
| W | * | * | * | * |
| X | 0.210 | 0.178 | 1.18 (0.90, 1.49) | 0.033 (-0.019, 0.076) |
| Y | 0.162 | 0.119 | 1.37 (1.25, 1.48) | 0.044 (0.032, 0.055) |
| Z | * | * | * | * |
| AA | 0.177 | 0.123 | 1.44 (1.23, 1.70) | 0.054 (0.030, 0.079) |
| BB | 0.272 | 0.165 | 1.65 (1.26, 2.30) | 0.107 (0.049, 0.173) |
| CC | 0.152 | 0.106 | 1.43 (1.30, 1.57) | 0.046 (0.034, 0.058) |
| DD | 0.183 | 0.144 | 1.27 (1.16, 1.40) | 0.039 (0.024, 0.054) |
| EE | * | * | * | * |
| FF | 0.185 | 0.152 | 1.22 (0.88, 1.73) | 0.033 (-0.022, 0.085) |
| GG | * | * | * | * |
| HH | * | * | * | * |
| II | 0.172 | 0.131 | 1.31 (1.17, 1.49) | 0.041 (0.023, 0.059) |
| JJ | 0.123 | 0.065 | 1.88 (1.03, 3.37) | 0.058 (0.003, 0.105) |
| KK | 0.141 | 0.111 | 1.28 (1.00, 1.69) | 0.031 (0.000, 0.065) |
| LL | 0.146 | 0.118 | 1.23 (1.07, 1.42) | 0.027 (0.009, 0.047) |
| MM | 0.087 | 0.027 | 3.19 (1.63, 10.28) | 0.060 (0.025, 0.106) |
| NN | 0.138 | 0.079 | 1.75 (1.09, 2.89) | 0.059 (0.008, 0.108) |
| OO | 0.194 | 0.142 | 1.37 (1.21, 1.54) | 0.053 (0.031, 0.072) |
| PP | 0.234 | 0.143 | 1.64 (1.36, 2.00) | 0.092 (0.056, 0.123) |
| QQ | 0.107 | 0.069 | 1.55 (1.19, 2.03) | 0.038 (0.016, 0.060) |
| RR | * | * | * | * |

eTable 2. Matched cohort description for the vaccine subanalysis; reinfection and control cohorts are perfectly balanced across all descriptors except the long COVID outcome. All cells have been perturbed by up to ±5 and totals recalculated to preserve obfuscation of counts <20.

| Characteristic | Total (N=168,146) | No Vaccinations (N=168,146) | Only Before First (N=168,146) | Only Before Index (N=168,146) | Before First and Index (N=168,146) |
|---|---|---|---|---|---|
| Long COVID | | | | | |
| Among Reinfection Cases | 6,533 (3.8%) | 2,491 (2.8%) | 1,435 (5.4%) | 2,052 (4.5%) | 555 (7.1%) |
| Among Control Cases | 4,943 (2.9%) | 1,769 (2.0%) | 1,028 (3.9%) | 1,676 (3.7%) | 470 (6.0%) |
| Sex | | | | | |
| Female | 105,858 (62.3%) | 53,096 (58.8%) | 18,093 (68.5%) | 29,747 (65.6%) | 4,922 (63.0%) |
| Male | 63,991 (37.7%) | 37,192 (41.2%) | 8,306 (31.5%) | 15,598 (34.4%) | 2,895 (37.0%) |
| Age (in years) | | | | | |
| 18-25 | 30,519 (18.0%) | 20,166 (22.3%) | 3,688 (14.0%) | 6,376 (14.1%) | 289 (3.7%) |
| 26-35 | 37,526 (22.1%) | 24,815 (27.5%) | 5,249 (19.9%) | 6,816 (15.0%) | 646 (8.3%) |
| 36-50 | 45,257 (26.6%) | 24,290 (26.9%) | 7,316 (27.7%) | 12,287 (27.1%) | 1,364 (17.4%) |
| 51-65 | 34,526 (20.3%) | 14,336 (15.9%) | 5,753 (21.8%) | 12,150 (26.8%) | 2,287 (29.3%) |
| 66+ | 22,029 (13.0%) | 6,684 (7.4%) | 4,395 (16.6%) | 7,719 (17.0%) | 3,231 (41.3%) |
| Race and Ethnicity | | | | | |
| American Indian or Alaska Native Non-Hispanic | 296 (0.2%) | 144 (0.2%) | 85 (0.3%) | 51 (0.1%) | <=20 (-) |
| Asian Non-Hispanic | 473 (0.3%) | 57 (0.1%) | 210 (0.8%) | 110 (0.2%) | 96 (1.2%) |
| Black or African American Non-Hispanic | 18,738 (11.0%) | 10,484 (11.6%) | 2,750 (10.4%) | 4,680 (10.3%) | 824 (10.5%) |
| Hispanic or Latino Any Race | 11,363 (6.7%) | 5,893 (6.5%) | 1,371 (5.2%) | 3,819 (8.4%) | 280 (3.6%) |
| Native Hawaiian or Other Pacific Islander Non-Hispanic | <=20 (-) | <=20 (-) | <=20 (-) | <=20 (-) | <=20 (-) |
| Other Non-Hispanic | 6,682 (3.9%) | 4,994 (5.5%) | 612 (2.3%) | 950 (2.1%) | 126 (1.6%) |
| Unknown | 2,317 (1.4%) | 614 (0.7%) | 408 (1.5%) | 1,207 (2.7%) | 88 (1.1%) |
| White Non-Hispanic | 129,989 (76.5%) | 68,120 (75.4%) | 20,967 (79.4%) | 34,522 (76.1%) | 6,380 (81.7%) |
| Charlson Comorbidity Index | | | | | |
| 0 | 138,363 (81.5%) | 78,506 (86.9%) | 19,303 (73.1%) | 35,839 (79.0%) | 4,715 (60.3%) |
| 1-2 | 23,457 (13.8%) | 9,502 (10.5%) | 5,003 (19.0%) | 6,951 (15.3%) | 2,001 (25.6%) |
| 3-4 | 5,480 (3.2%) | 1,644 (1.8%) | 1,326 (5.0%) | 1,750 (3.9%) | 760 (9.7%) |
| 5+ | 2,562 (1.5%) | 646 (0.7%) | 768 (2.9%) | 805 (1.8%) | 343 (4.4%) |
| Number of Visit Days in Year Before First COVID-19 Event | | | | | |
| 0 | 19,633 (11.6%) | 15,078 (16.7%) | 1,215 (4.6%) | 3,124 (6.9%) | 216 (2.8%) |
| 1-2 | 25,802 (15.2%) | 17,116 (19.0%) | 2,355 (8.9%) | 5,902 (13.0%) | 429 (5.5%) |
| 3-5 | 26,176 (15.4%) | 15,070 (16.7%) | 3,396 (12.9%) | 7,034 (15.5%) | 676 (8.7%) |
| 6-12 | 35,757 (21.1%) | 17,607 (19.5%) | 5,889 (22.3%) | 10,820 (23.9%) | 1,441 (18.5%) |
| 13-23 | 27,727 (16.3%) | 11,884 (13.2%) | 5,315 (20.1%) | 8,782 (19.4%) | 1,746 (22.4%) |
| 24-54 | 24,127 (14.2%) | 9,483 (10.5%) | 5,497 (20.8%) | 6,960 (15.3%) | 2,187 (28.0%) |
| 55+ | 10,614 (6.2%) | 4,051 (4.5%) | 2,727 (10.3%) | 2,730 (6.0%) | 1,106 (14.2%) |
| First Infection Severity | | | | | |
| No Hospitalization or Emergency Department | 141,746 (83.5%) | 72,739 (80.6%) | 22,487 (85.1%) | 39,679 (87.5%) | 6,841 (87.6%) |
| Emergency Department with No Hospitalization | 21,807 (12.8%) | 14,329 (15.9%) | 2,957 (11.2%) | 3,872 (8.5%) | 649 (8.3%) |

| | | | | | |
|---|---|---|---|---|---|
| Moderate Hospitalization | 6,270 (3.7%) | 3,209 (3.6%) | 957 (3.6%) | 1,785 (3.9%) | 319 (4.1%) |
| Severe Hospitalization with Ventilation or ECMO | 25 (0.0%) | <=20 (-) | <=20 (-) | <=20 (-) | <=20 (-) |
| Quarter of First COVID-19 Event | | | | | |
| 2020 Quarter 1 | 263 (0.2%) | 79 (0.1%) | <=20 (-) | 187 (0.4%) | <=20 (-) |
| 2020 Quarter 2 | 4,134 (2.4%) | 1,835 (2.0%) | <=20 (-) | 2,297 (5.1%) | <=20 (-) |
| 2020 Quarter 3 | 8,093 (4.8%) | 3,278 (3.6%) | <=20 (-) | 4,816 (10.6%) | <=20 (-) |
| 2020 Quarter 4 | 43,377 (25.5%) | 16,876 (18.7%) | <=20 (-) | 26,508 (58.5%) | <=20 (-) |
| 2021 Quarter 1 | 16,780 (9.9%) | 7,187 (8.0%) | 27 (0.1%) | 9,244 (20.4%) | 322 (4.1%) |
| 2021 Quarter 2 | 6,693 (3.9%) | 4,866 (5.4%) | 118 (0.4%) | 1,336 (2.9%) | 373 (4.8%) |
| 2021 Quarter 3 | 22,203 (13.1%) | 17,072 (18.9%) | 1,988 (7.5%) | 679 (1.5%) | 2,464 (31.6%) |
| 2021 Quarter 4 | 27,047 (15.9%) | 19,571 (21.7%) | 5,721 (21.7%) | 202 (0.4%) | 1,553 (19.9%) |
| 2022 Quarter 1 | 26,192 (15.4%) | 14,843 (16.4%) | 9,970 (37.8%) | 64 (0.1%) | 1,315 (16.9%) |
| 2022 Quarter 2 | 5,227 (3.1%) | 1,621 (1.8%) | 2,612 (9.9%) | <=20 (-) | 986 (12.6%) |
| 2022 Quarter 3 | 5,712 (3.4%) | 1,957 (2.2%) | 3,013 (11.4%) | <=20 (-) | 740 (9.5%) |
| 2022 Quarter 4 | 2,696 (1.6%) | 781 (0.9%) | 1,881 (7.1%) | <=20 (-) | 31 (0.4%) |
| 2023 Quarter 1 | 1,054 (0.6%) | 247 (0.3%) | 798 (3.0%) | <=20 (-) | <=20 (-) |
| 2023 Quarter 2 | 200 (0.1%) | 38 (0.0%) | 161 (0.6%) | <=20 (-) | <=20 (-) |
| 2023 Quarter 3 | 125 (0.1%) | 23 (0.0%) | 103 (0.4%) | <=20 (-) | <=20 (-) |
